# A Match Made in Cyberspace: Applicant Perspectives on a Virtual Residency Application Process

**DOI:** 10.1101/2022.02.07.22270645

**Authors:** Shirley Chen, Melanie Schroeder, Thomas Kun Pak, Natasha Topolski, Emelyn Zaworski, Kseniya Anishchenko, Shyon Parsa, Christina Zhu, Esther Bae, Whitney Stuard, Jay Patel, Shanon Quach, Sahar Panjwani

**Affiliations:** Vanderbilt University School of Medicine, Nashville, Tennessee; University of Arizona College of Medicine - Phoenix, Phoenix, Arizona; University of Iowa, Iowa City, Iowa; McGovern Medical School at University of Texas Health Science Center, Houston, Texas; Medical College of Wisconsin, Wauwatosa, Wisconsin; University of Colorado School of Medicine, Aurora, Colorado; University of Texas Southwestern Medical School, Dallas, Texas; Texas Tech University Health Science Center School of Medicine, Lubbock, Texas; Wayne State University School of Medicine, Detroit, Michigan; University of Tennessee Health Science Center College of Medicine, Memphis, Tennessee; University of North Texas Health Science Center, Texas College of Osteopathic Medicine, Fort Worth, Texas; University of Texas Rio Grande Valley School of Medicine, Edinburg, Texas

## Abstract

**Background:** The COVID-19 pandemic precipitated an entirely virtual 2020-2021 residency application cycle. With many specialties continuing virtual interviews, it remains essential to evaluate all stakeholders perspectives.

**Objective:** This study aimed to evaluate applicants’ perspectives on the completely virtual 2020-2021 Match.

**Methods:** An online survey utilizing Likert scale and free response questions was distributed to residency applicants in 2021 post-Match to query their perspectives on the virtual application cycle.

**Results:** Participants (n=158/2811, 5% estimated response rate) represented 24 states and applied to 31 specialties. Most were satisfied with their Match experience (73.1%) and thought virtual interviews should continue (68.6%). However, applicants felt that the virtual setting permitted interview hoarding (73.9%) and that their medical school did not provide adequate electronic equipment (66.1%). Applicants found Twitter (78.4%) and Instagram (69.1%) to be helpful social media platforms for evaluating fit. While many applicants (56.9%) did not support application limits, most supported interview limits (62.7%) and preference signaling mechanisms (59.7%). Free responses data elucidated applicants’ views on how the virtual Match impacted financial access, evaluation of program culture, and the future of residency interviews.

**Conclusions:** Our study highlights that, across geography and specialty, many applicants support virtual interviews continuing. It also offers insight into how medical schools and residency programs can support applicants in the virtual environment. Additionally, the virtual setting provides an opportunity to evaluate mechanisms for addressing congestion within the Match, with most applicants supporting interview caps and preference signaling.

## Introduction

The COVID-19 pandemic caused a paradigm shift in the residency application process, with precautions requiring the suspension of in-person away rotations and pivoting to virtual interviews during the 2020-2021 cycle.^1–3^

Existing studies on the 2020-2021 virtual application process surveyed applicants before interviews occurred, asked residents in the context of fellowship applications, or assessed how faculty viewed virtual interviews.^4–10^ While studies indicate that fellowship program directors prefer in-person interviews, residents applying for fellowship expressed mixed opinions about whether virtual interviews should continue.^5–10^ Fellowship applicants noted the financial and time benefits of virtual interviews but expressed concerns with evaluating fit.^5–8^ A comprehensive evaluation of a virtual Match process requires input from all stakeholders, including applicants to residency positions. The data remain unclear regarding whether this population wants virtual interviews to continue.^4,5,8^

One noted benefit of virtual interviews is the greatly reduced monetary commitment. Before the pandemic, there were calls for change regarding the financial costs of residency applications, with several studies finding that video interviews could be a cost-saving alternative to traditional in-person interviews.^11–15^ The 2020-2021 Match confirmed this, as the virtual application process was estimated to save candidates nearly $6,000.^16^ The switch to virtual interviews also sparked a surge in residency programs’ social media presence.^17–20^ However, the utility of social media and other remote resources such as websites and virtual programming for applicants remains unclear. Another area of study is evaluating policies that address the increasing number of applications and interviews per applicant.^21^ Finally, virtual interviews may exacerbate inequities for those without access to electronic equipment and high-speed Internet, a phenomenon termed digital redlining.^22–24^

Our study aims to evaluate applicants’ perspectives on the 2020-2021 virtual residency application cycle. We identified three key outcomes: [1] general thoughts including overall satisfaction and concerns with the virtual environment, [2] utility of resources provided by programs for evaluating fit, and [3] perspectives on changes to make during future cycles.

## Methods

This study aimed to characterize applicants’ perspectives post-Match on the impact of a completely virtual setting on the 2020-2021 residency application process. A survey was distributed through a national medical student organization email list and grassroots social media efforts.

Given that the 2020-2021 cycle was the first completely virtual residency application process, no prior surveys were available to assess student opinions on this topic. Therefore, a 30-question survey instrument was developed by consulting members of a national medical student committee focused on medical education, 2020-2021 residency applicants, and survey methodology experts. Surveys by the National Resident Matching Program and specific specialties were also referenced.

Five M.D. and/or Ph.D. methodology experts evaluated the appropriateness of our instrument in addressing the study’s objectives as part of pre-testing. A pilot study was then conducted with 20 fourth-year medical students to ensure question clarity prior to wide distribution.

We identified three key outcomes to assess: [1] general thoughts, [2] program resources, and [3] perspectives on future cycles. First, it was important to characterize applicants’ overall satisfaction with the cycle and concerns unique to the virtual setting. Second, given how applicants were not able to visit programs in-person, we aimed to identify what resources provided by residencies were most useful for determining compatibility with a program. Third, we assessed applicant opinions on policies to implement during future cycles in response to the virtual setting.

Most questions utilized a 5-point Likert scale by which students either rated their agreement with statements about the application cycle or the helpfulness of listed resources. Five free response questions allowed applicants to elaborate on their application experience. The survey instrument can be accessed in the online supplemental content.

Response percentages, 95% confidence intervals (CIs), and statistical significance were calculated using a customized Python computing package.^25^ For analysis of survey items using Likert scales, categories of “Agree” and “Strongly Agree” were grouped together as “Agree”, “Disagree” and “Strongly Disagree” were grouped together as “Disagree”, “Helpful” and “Extremely Helpful” were grouped together as “Helpful”, and “Unhelpful” and “Extremely Unhelpful” were grouped together as “Unhelpful”.^26^ Statistical significance was determined by testing for overlap among the 95% CIs for “Agree”, “Neutral”, and “Disagree” for questions asking about agreement with a statement and “Helpful”, “Neutral”, and “Unhelpful” for questions asking about the helpfulness of a resource. For analysis of free response questions, three independent reviewers identified common themes that provided nuance to the Likert scale results. Discrepancies were resolved by discussion among the analysts.

The Vanderbilt Human Research Protections Program granted this study Institutional Review Board exemption.

## Results

The survey was emailed to members of a national medical student organization. 11,243 students viewed the correspondence and, of those, we estimate that 25% were eligible to participate since students apply to residency during their last year of medical school. Therefore, we conclude that 2,811 email recipients met study criteria. Given that grassroots social media efforts were also utilized, students contacted via social media likely overlapped with students contacted via the national organization. It is therefore not possible to determine the exact numbers of students reached with these combined methods. With a total of 158 responses, we conservatively estimate our response rate to be approximately 5%. This survey captured individuals from 24 states who applied into 31 specialties, including 8 combined specialty programs (Table 1).

**Table 1:**
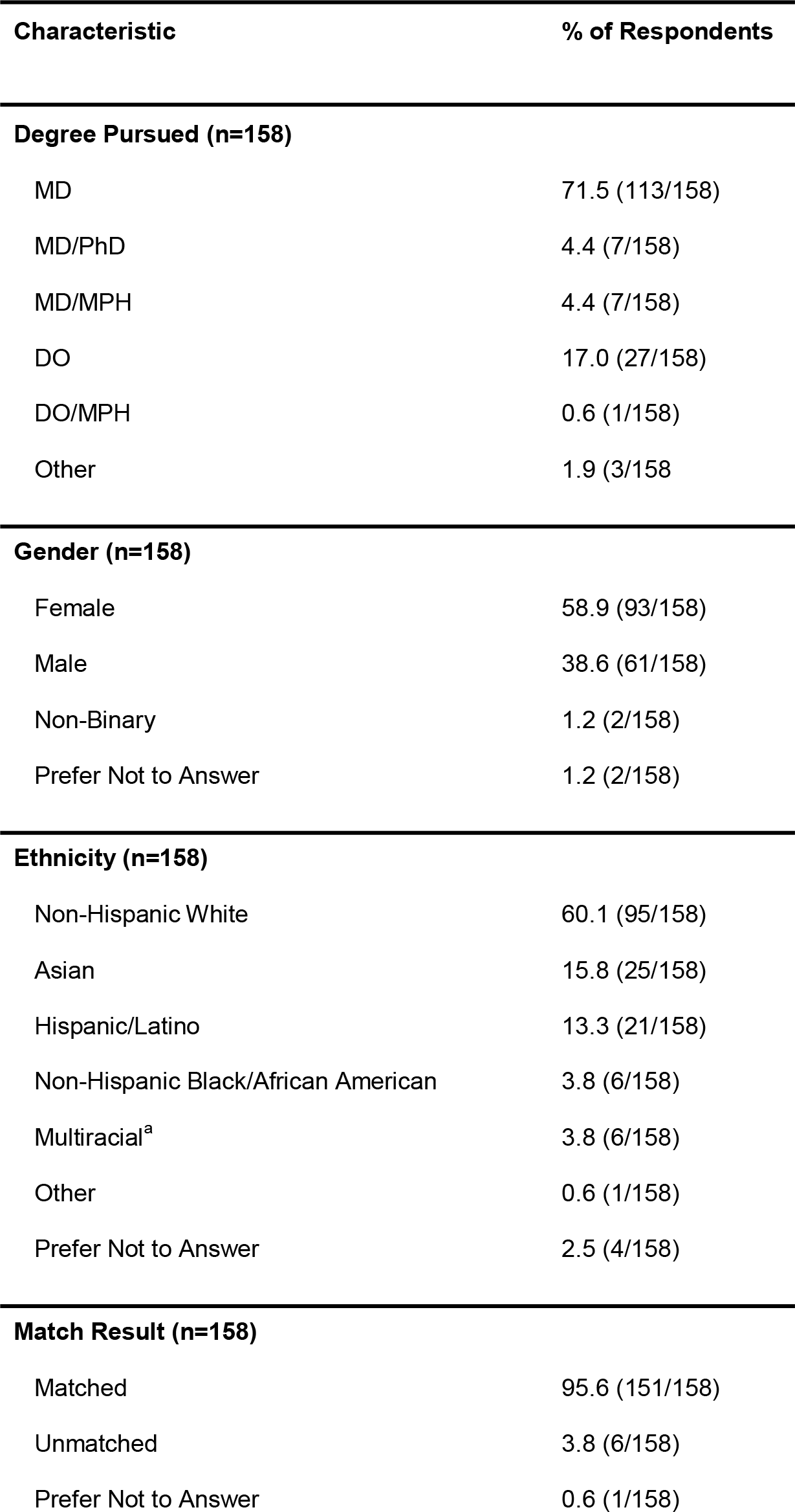
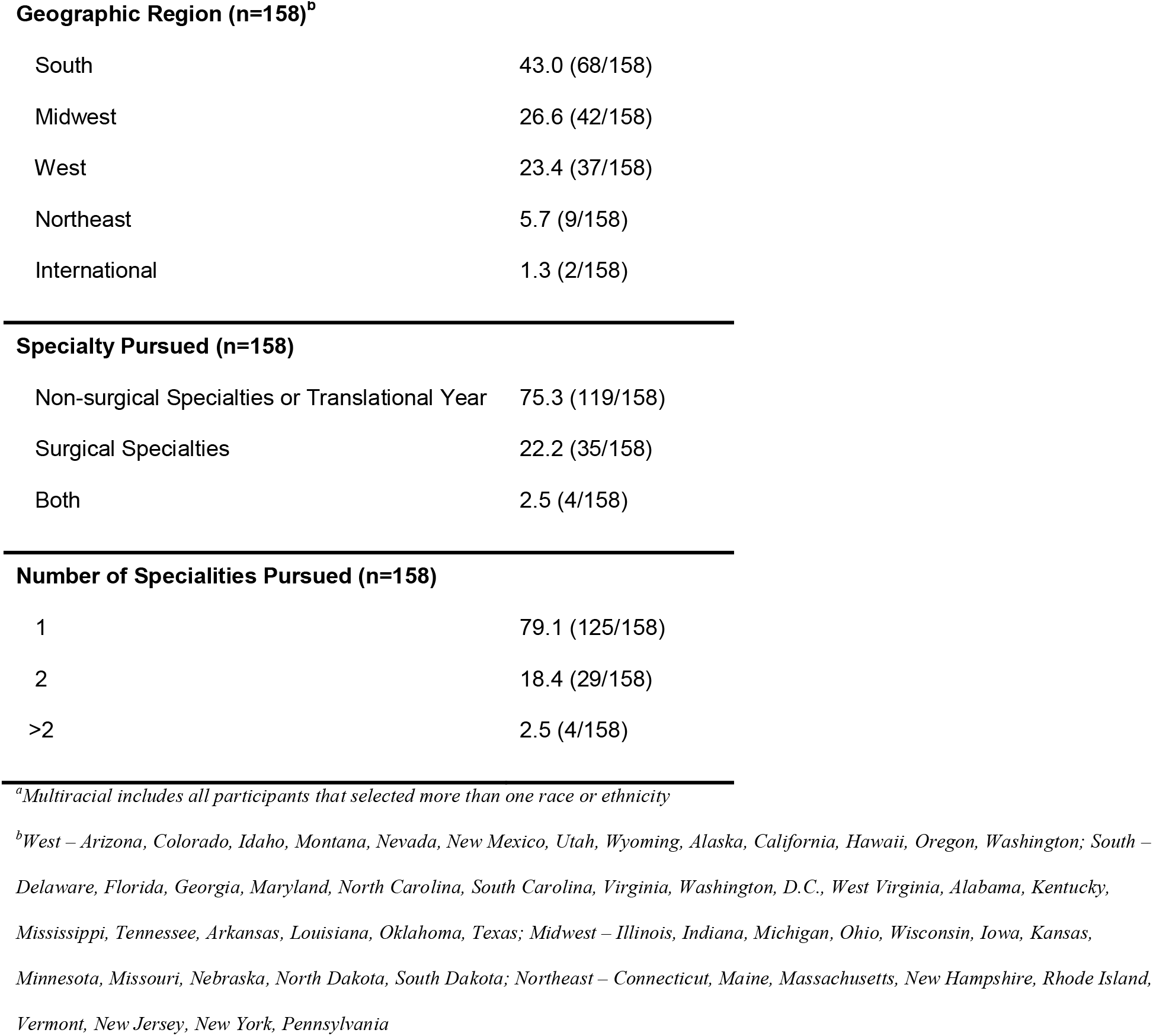
Characteristics of survey respondents.

| Characteristic | % of Respondents |
| --- | --- |
| **Degree Pursued (n=158)** |  |
| MD | 71.5 (113/158) |
| MD/PhD | 4.4 (7/158) |
| MD/MPH | 4.4 (7/158) |
| DO | 17.0 (27/158) |
| DO/MPH | 0.6 (1/158) |
| Other | 1.9 (3/158) |
| **Gender (n=158)** |  |
| Female | 58.9 (93/158) |
| Male | 38.6 (61/158) |
| Non-Binary | 1.2 (2/158) |
| Prefer Not to Answer | 1.2 (2/158) |
| **Ethnicity (n=158)** |  |
| Non-Hispanic White | 60.1 (95/158) |
| Asian | 15.8 (25/158) |
| Hispanic/Latino | 13.3 (21/158) |
| Non-Hispanic Black/African American | 3.8 (6/158) |
| Multiracial<sup>a</sup> | 3.8 (6/158) |
| Other | 0.6 (1/158) |
| Prefer Not to Answer | 2.5 (4/158) |
| **Match Result (n=158)** |  |
| Matched | 95.6 (151/158) |
| Unmatched | 3.8 (6/158) |
| Prefer Not to Answer | 0.6 (1/158) |
| **Geographic Region (n=158)<sup>b</sup>** |  |
| South | 43.0 (68/158) |
| Midwest | 26.6 (42/158) |
| West | 23.4 (37/158) |
| Northeast | 5.7 (9/158) |
| International | 1.3 (2/158) |
| **Specialty Pursued (n=158)** |  |
| Non-surgical Specialties or Translational Year | 75.3 (119/158) |
| Surgical Specialties | 22.2 (35/158) |
| Both | 2.5 (4/158) |
| **Number of Specialities Pursued (n=158)** |  |
| 1 | 79.1 (125/158) |
| 2 | 18.4 (29/158) |
| >2 | 2.5 (4/158) |

<sup>a</sup>Multiracial includes all participants that selected more than one race or ethnicity
<sup>b</sup>West – Arizona, Colorado, Idaho, Montana, Nevada, New Mexico, Utah, Wyoming, Alaska, California, Hawaii, Oregon, Washington; South – Delaware, Florida, Georgia, Maryland, North Carolina, South Carolina, Virginia, Washington, D.C., West Virginia, Alabama, Kentucky, Mississippi, Tennessee, Arkansas, Louisiana, Oklahoma, Texas; Midwest – Illinois, Indiana, Michigan, Ohio, Wisconsin, Iowa, Kansas, Minnesota, Missouri, Nebraska, North Dakota, South Dakota; Northeast – Connecticut, Maine, Massachusetts, New Hampshire, Rhode Island, Vermont, New Jersey, New York, Pennsylvania

### Benefits and concerns with a virtual residency application process

The majority of respondents were satisfied with their 2020-2021 Match experience (114 of 156 [73.1%]; P<0.05; 95% CI 66.1-80.0; Table 2). When asked if they thought the virtual setting allowed more students to hoard interviews, a significant proportion of respondents agreed (116 of 157 [73.9%]; P<0.05; 95% CI 67.0-80.8; Table 2). Most respondents agreed that the lack of in-person away rotations limited their ability to evaluate programs (81 of 157 [51.6%]; P<0.05; 95% CI 43.8-59.4; Table 2). Furthermore, ranking residency programs was a concern, with a majority agreeing that the virtual setting made the ranking process challenging (98 of 157 [62.4%]; P<0.05; 95% CI 54.8-70.0; Table 2).

**Table 2:** General applicant perspectives on a virtual Match^a^.

| **Statement** | **% Disagree,**<br>**[95% CI]** | **% Neutral,**<br>**[95% CI]** | **% Agree,**<br>**[95% CI]** |
| --- | --- | --- | --- |
| **Overall, I am satisfied with my residency application/virtual interview experience this year. (n=156)** | 18.6 (29/156),<br>[12.5-24.7] | 8.3 (13/156),<br>[4.0-12.7] | **73.1 (114/156)**<br>**[66.1-80.0]** |
| **I feel that the virtual setting allowed more students to 'hoard' interviews. (n=157)** | 9.6 (15/157),<br>[5.0-14.2] | 16.6 (26/157),<br>[10.7-22.4] | **73.9 (116/157),**<br>**[67.0-80.8]** |
| **The virtual setting made it difficult to be myself when interacting with programs. (n=157)** | 44.6 (70/157),<br>[36.8-52.4] | 18.5 (29/157),<br>[12.4-24.5] | 36.9 (58/157),<br>[29.4-44.5] |
| **The virtual setting decreases an applicant's ability to show genuine interest in a program. (n=154)** | 35.1 (54/154),<br>[27.5-42.6] | 16.2 (25/154),<br>[10.4-22.1] | 48.7 (75/154),<br>[40.8-56.6] |
| **The lack of in-person away rotations limited my ability to assess residency programs. (n=157)** | 26.1 (41/157),<br>[19.2- 33.0] | 22.3 (35/157),<br>[15.8-28.8] | **51.6 (81/157),**<br>**[43.8-59.4]** |
| **The virtual setting allowed me to effectively evaluate my 'fit' within a program. (n=157)** | 36.9 (58/157),<br>[29.4-44.5] | 22.9 (36/157),<br>[16.4-29.5] | 40.1 (63/157),<br>[32.5-47.8] |
| **The virtual setting made the ranking process challenging. (n=157)** | 23.6 (37/157)<br>[16.9-30.2] | 14.0 (22/157),<br>[8.6-19.4] | **62.4 (98/157),**<br>**[54.8-70.0]** |
| **The virtual setting increases financial access to interviews. (n=157)** | X<sup>b</sup> | 1.9 (3/157),<br>[0-4.1] | **98.1 (154/157),**<br>**[95.9-100]** |
| The virtual setting allowed me to attend more interviews because of reduced financial constraints. (n=157) | 6.4 (10/157),<br>[2.5-10.2] | 15.9 (25/157),<br>[10.2-21.6] | **77.7 (122/157),**<br>**[71.2-84.2]** |
| I felt that I had the resources to successfully complete my residency interviews. (n=155) | 5.8 (9/155),<br>[2.1-9.5] | 9.0 (14/155),<br>[4.5-13.5] | **85.2 (132/155),**<br>**[79.6-90.8]** |
| My school provided adequate physical space for virtual interviews. (n=132) | 30.3 (40/132),<br>[22.5-38.1] | 9.8 (13/132),<br>[4.8-14.9] | **59.8 (79/132),**<br>**[51.5-68.2]** |
| My school provided adequate coaching and/or guidance on how to prepare for virtual interviews. (n=150) | 18.7 (28/150),<br>[12.4-24.9] | 18.0 (27/150),<br>[11.9-24.1] | **63.3 (95/150),**<br>**[55.6-71.0]** |
| My school provided adequate electronic equipment (webcams, lights, etc.) for virtual interviews. (n=118) | **66.1 (78/118),**<br>**[57.6-74.6]** | 12.7 (15/118),<br>[6.7-18.7] | 21.2 (25/118),<br>[13.8-28.6] |

<sup>a</sup>Bolded text indicates statistically significant result ( $P < 0.05$ )
<sup>b</sup>0 responses for this answer choice

Nearly all respondents agreed that the virtual setting increased financial access to interviews (151 of 154 [98.1%]; P<0.05; 95% CI 95.9-100; Table 2), and the majority agreed that they were able to attend more interviews because of reduced financial constraints (122 of 157 [77.7%]; P<0.05; 95% CI 71.2-84.2; Table 2). Beyond finances, most respondents agreed that they had the necessary resources to complete residency interviews (132 of 155 [85.2%]; P<0.05; 95% CI 79.6-90.8; Table 2). However, while a majority agreed that their school provided sufficient physical space for virtual interviews (79 of 132 [59.8%]; P<0.05; 95% CI 51.5-68.2; Table 2) and sufficient guidance on how to interview virtually (95 of 150 [63.3%]; P<0.05; 95% CI 55.6-71.0; Table 2), most respondents did not believe that their medical school provided adequate electronic equipment for virtual interviews (78 of 118 [66.1%]; P< 0.05; 95% CI 57.6-74.6; Table 2).

### Utility of resources provided by residency programs in evaluating fit

When assessing compatibility with a program, the majority of respondents agreed that Twitter (58 of 74 [78.4%]; P<0.05%; 95% CI 69.0-87.8; Table 3) and Instagram (76 of 110 [69.1%]; P<0.05; 95% CI 60.5-77.7; Table 3) were helpful. Conversely, a majority thought that Tik Tok was unhelpful (12 of 17 [70.6%]; P<0.05; 95% CI 48.9-92.2; Table 3)

**Table 3:**
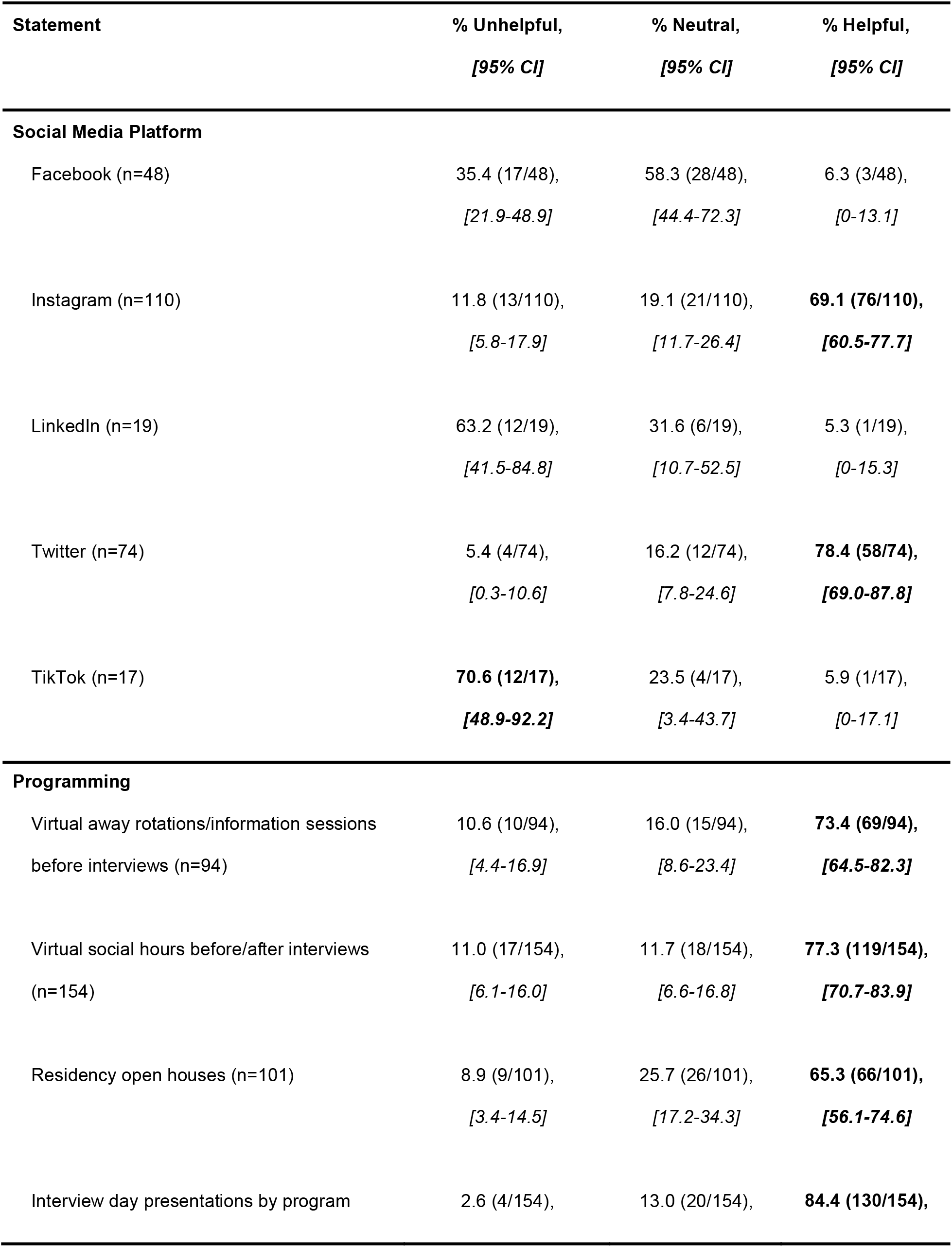
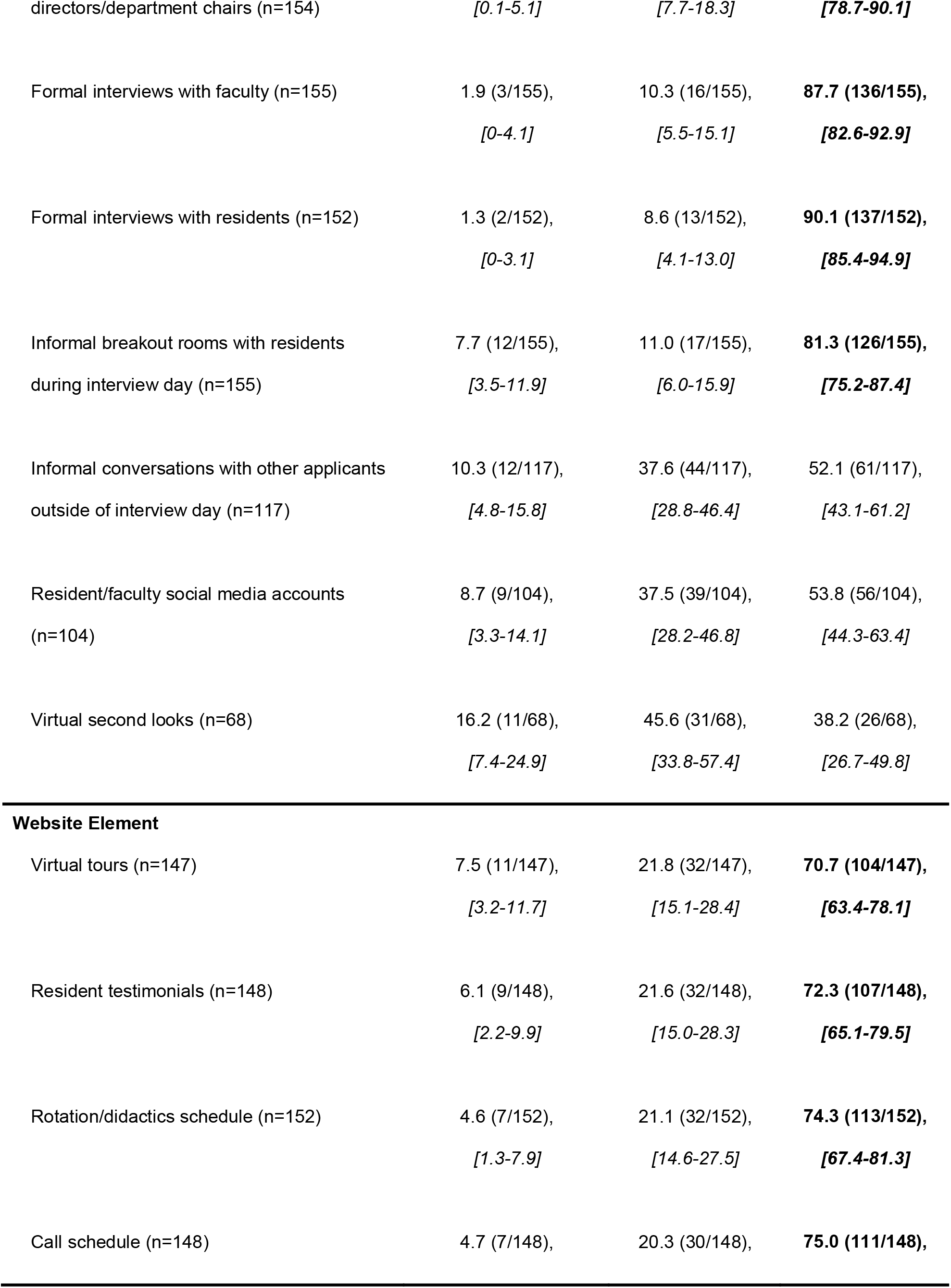
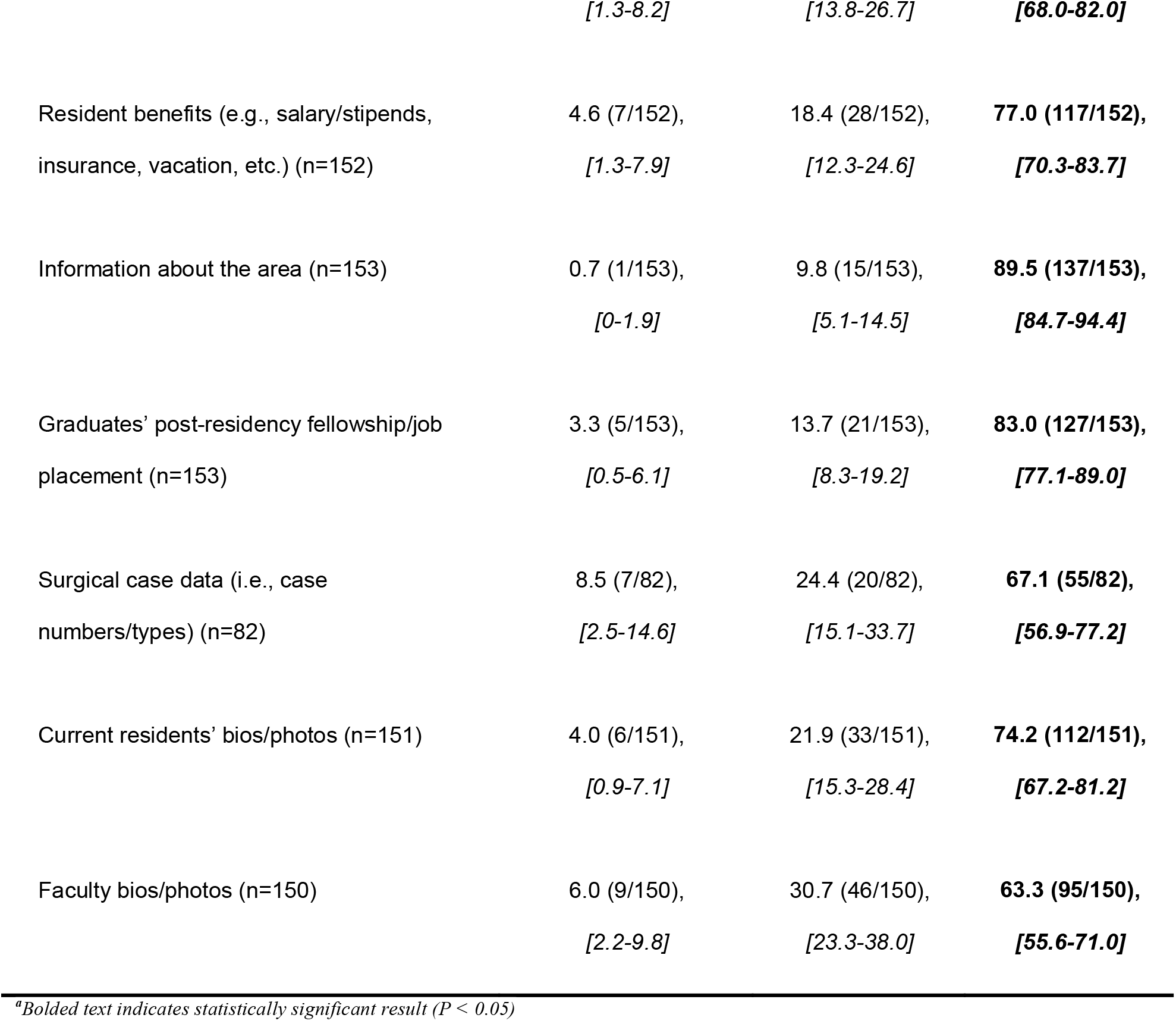
Applicant perspectives on helpfulness of residency program resources^a^.

Interestingly, while respondents agreed that all aspects of program websites were helpful and most programming organized by residencies were helpful, virtual second looks (26 of 68 [38.2%]; P>0.05; 95% CI 26.7-49.8; Table 3), informal conversations with other applicants (61 of 117 [52.1%]; P>0.05; 95% CI 43.1-61.2; Table 3), and resident or faculty social media accounts (56 of 104 [53.8%]; P>0.05; 95% CI 44.3-63.4; Table 3) were not found to be significantly helpful by respondents.

### Changes to the residency application process

When asked about future application cycles, the majority of respondents did not support an application limit (87 of 153 [56.9%]; P<0.05; 95% CI 49.0-64.7; Table 4) but supported an interview limit (96 of 153 [62.7%]; P<0.05; 95% CI 55.1-70.4; Table 4). Most also agreed with implementing a preference signaling mechanism to indicate genuine interest in a program (89 of 149 [59.7%]; P<0.05; 95% CI 51.9-67.6; Table 4). Finally, the majority of respondents thought that residency programs should continue to provide virtual interviews even when in-person interviews are again possible (105 of 153 [68.6%]; P<0.05; CI 61.3-76.0; Table 4).

**Table 4:**
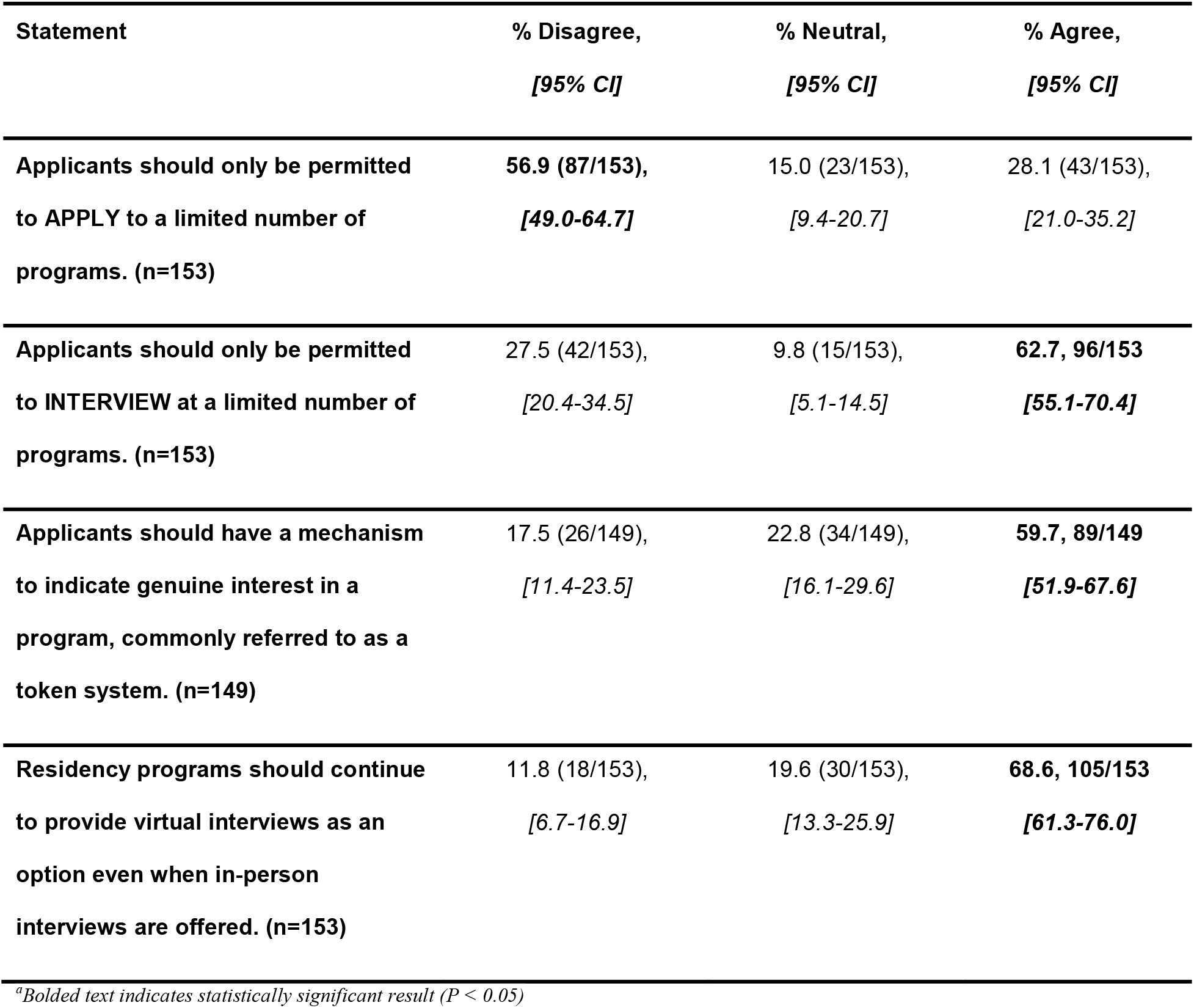
Applicant perspectives on future residency application cycles^a^.

### Additional insights from free response questions

The free response comments add nuance to the Likert scale findings outlined above. Five major themes emerged: [1] Financial access: while virtual interviews can improve equity by increasing financial access, respondents expressed that finances should not be the only consideration when deciding whether to continue virtual interviews. [2] Interview resources: access to resources varied greatly among participants, and provision of resources by medical schools—including physical space, electronic equipment, and preparation tools such as third party mock interview software—were highly valued by applicants. [3] Online presence: a residency’s online presence greatly impacted applicants’ perception of the program, but concerns existed about the role of social media in future cycles, including the possibility of students being required to utilize social media during this process and programs evaluating applicants by their social media presence. [4] Evaluation of program culture: several factors hindered applicants’ ability to evaluate fit with a program, including a lack of meaningful interactions with residents and faculty, an inability to observe the hospital environment, and the possibility of programs curating their online presence. [5] Future of residency interviews: respondents had conflicting opinions about the future of residency interviews, with many concerned about equity regarding optional virtual interviews. Additional insights and illustrative quotes can be found in Supplemental Tables 1-5.

## Discussion

Most applicants were satisfied with the 2020-2021 Match and wanted residencies to continue providing virtual interviews. However, many expressed equity concerns including greater opportunities for interview hoarding and disparities in resources provided by medical schools. For determining fit, Instagram and Twitter were found to be particularly helpful social media platforms, and information on official program websites was consistently found to be helpful. The majority of applicants supported a preference signaling mechanism and interview limit, but only a minority supported an application limit.

Pre-pandemic studies disagreed on whether applicants preferred virtual interviews to the traditional in-person format.^27,28^ Those supporting virtual interviews primarily did so due to financial considerations.^28^ Our survey of applicants who participated in a completely virtual residency application process demonstrated that the majority were satisfied with their Match experience and want virtual interviews to continue. However, free response comments indicated equity concerns if virtual interviews are offered as an alternative to in-person interviews due to the travel costs associated with the in-person format. Although our study was concordant with past studies finding that most applicants felt they had adequate resources to complete virtual interviews, a majority of our participants thought that their schools did not provide sufficient electronic resources.^2^ As virtual interviews continue, this could be an opportunity for medical schools to combat digital redlining by providing students with the technology required to succeed in the virtual setting.^29–31^ A recognized disadvantage of virtual interviews is the difficulty applicants have with evaluating compatibility with a program.^27^ This is evidenced by how most of our study participants agreed that a completely virtual application process made ranking programs more challenging. To address the challenge of evaluating fit virtually, applicants have increasingly relied on the social media presence of programs.^32,33^ Few studies have evaluated the utility of social media platforms for applicants in this context. Interestingly, one pre-pandemic study found that students did not believe Twitter or Instagram presence to be important when evaluating residency programs.^27^ Our results suggest that not only did study participants find Instagram and Twitter to be helpful when evaluating program compatibility, they also found websites on the whole and most programming organized by residencies to be helpful. However, free response comments expressed caution with this increased reliance on social media due to the potential for developing bias towards students more active on social media.

Our study found that the abrupt adoption of virtual interviews raised other concerns. One was students’ increased ability to hoard interviews in the virtual setting. Possible means to address this include application and interview limits, with our study only demonstrating majority support for the latter. Interestingly, a few specialties—emergency medicine, ophthalmology, and psychiatry— implemented interview caps for the 2021-2022 cycle.^34,35^ Additionally, most of our study participants supported preference signaling to address the congestion associated with the increasing number of applications residency programs receive.^21^ Otolaryngology implemented a preference signaling system during the 2020-2021 Match, and the majority of the participants were satisfied with their experience.^36^ During the 2021-2022 cycle, more specialties—internal medicine, dermatology, and general surgery—are utilizing a preference signaling mechanism within their supplemental applications.^37^ With more widespread adoption of both interview limits and preference signaling, it remains to be seen how these new policies will be received by Match stakeholders. Considering these findings and results from other studies, we propose four recommendations (Figure 1).^2,27,28,30,31,34–41^

Our study has several limitations. Given that survey participation was voluntary, there was potential for response bias towards applicants with particularly strong feelings about their Match experience. Despite extensive grassroots efforts, we collected responses from only a small fraction of all Match participants, limiting analysis based on applicant identity, geography, and specialty. Additionally, since all survey items were optional, there were variable response rates across questions. Due to the lack of standardized survey instruments on this topic, we developed our own survey questions and are only able to demonstrate internal validity through pilot testing. Further research on this topic with larger sample sizes or qualitative methodology is needed to solidify students’ perspectives on this topic

Future directions for our study include assessing applicant satisfaction with interview caps and preference signaling in the virtual setting. With the possibility of in-person interviews continuing, applicant satisfaction with a hybrid interview process should also be evaluated.

## Conclusions

Our study demonstrates that, across specialty and geography, Match participants support virtual interviews. Residencies can help applicants better evaluate program compatibility by investing in social media, official websites, and virtual programming, while medical schools can combat potential disparities by providing students with the resources needed for virtual interviews. Residency applicants also support interview caps and preference signaling to address the high number of applications residency programs receive and the greater potential to hoard interviews in the virtual setting.

## Supporting information

Survey Instrument

## Data Availability

All data produced in the present study are available upon reasonable request to the authors.

## Acknowledgements

The authors would like to thank Joseph Camerano, Rijul Asri, Ryan Chiu, Samuel Neher, Norman Farr, Matthew McEchron, Brian Drolet, and Christine Ford, and the other individuals who assisted with the development and distribution of this survey.

## Notes

### Competing Interest Statement

The authors have declared no competing interest.

### Funding Statement

This study did not receive any funding.

### Author Declarations

IRB #210572 "COVID Effects on Residency Applications" was approved for exemption status on 04/22/2021 by the Vanderbilt Human Research Protections Program - HRPP.

### Summary of Updates

Revised to be more concise and focused on significant results

