## Supplemental Tables for "A Match Made in Cyberspace: Applicant Perspectives on a Virtual Residency Application Process"

**Supplemental Table 1: Applicants’ Perspectives on Increased Financial Access with Virtual Interviews**

| **Financial Access**  While respondents agreed that virtual interviews can improve equity by increasing financial access, respondents also expressed that finances should not be the only consideration when determining the future of virtual interviews. | |
| --- | --- |
| **Sub-Theme** | **Representative Quotations** |
| **Access to Attend More Interviews** | - “Virtual interviews did a fantastic job at reducing financial barriers that many low-income students face. We have to dish out so much money to apply to, to stay in, and to thrive in medical school. Think about us for a minute. Think about those who cannot afford to purchase expensive airline tickets at the drop of a hat. Think about your parenting students who cannot simply leave their children and partners for weeks at a time. We still deserve the chance to be seen by our dream programs; reducing this barrier is critical to this.” - “The factors I considered when creating my rank list (mission, location, personality fit) were not limited by virtual interviews. Additionally, I feel much more financially stable for starting residency by not having spent so much money on travel.” |
| **Apprehension on Overweighting Cost as a Factor** | - “In-person interviews are far superior, not just for evaluating the programs but for getting to know the area that you might live in. Especially if you are moving with a partner and/or family, it is very difficult to base this crucial life decision via Zoom. There are financial benefits, however the cost of in-person interviews is a drop in the bucket compared to what we will earn as physicians in the years to come. I would strongly urge programs to return to in-person interviewing ASAP.” |

**Supplemental Table 2: Applicants’ Perspectives on Access to Resources in the Setting of Virtual Interviews**

| **Interview Resources**  While our survey indicated that the majority of applicants felt that their school provided adequate physical space, several respondents stated that they encountered difficulties, including space being limited, hard to book, or not available at needed times. Notably, although most respondents did not think they were provided adequate electronic equipment, students that did receive equipment remarked on the helpfulness of this resource along with training sessions on how to best utilize them. Moreover, one respondent mentioned how a student-led effort at their institution to provide resources was extremely helpful. Finally, while the majority of respondents indicated they received adequate coaching from their medical school on how to conduct virtual interviews, many comments indicated that the most useful resource provided by their institution was access to virtual mock interviews. Notably, several students mentioned third party interviewing resources such as Big Interview and Stand Out were particularly helpful. | |
| --- | --- |
| **Sub-Theme** | **Representative Quotations** |
| **Virtual Interview Preparation Resources Were Helpful** | - “My school paid for a subscription for us to practice answering interview questions, where you're recorded on your webcam and can re-watch your answers.” - “My school provided one 30-minute practice interview with a career-development team, which was immensely helpful and provided concrete feedback for improvement.” |
| **Physical Space Provided by Medical Schools Was Valued** | - “We were able to reserve very nice, single occupancy rooms in the student center to have a quiet and private place to interview. I believe there was an option for extra loan money for applications/interviews if needed.” - “Opened up rooms on campus for residency interviews. There was a fund we could apply to for equipment fundings” |
| **Issues with Space and Other Resources Needed to Conduct Virtual Interviews** | - “My school didn't provide anything, and because of COVID we weren't allowed to use any on-campus physical areas. When asked if we could, we were told it would be too challenging for them to figure out how to do so in a "safe manner". I had to interview in my parents house, in my old bedroom. Very very humbling.” - “My school gave us zero material resources and a very limited attempt at space for interviews (limited on-campus space that filled up with reservations quickly, could never realistically accommodate every student if they needed it), no webcams or lighting, etc. But [we] did get some counseling about interviews themselves.” - “Myself and other SGA [Student Government Association] reps organized virtual interview kits that could be checked out by students in the library. These kits included a laptop stand, ring light, USB port, webcam, pop-up photo background, and photo background stand. Students were expected to provide their own laptop. Students could check out rooms at the school to conduct interviews in. The list of items in the kit was also sent out to every student, so those students who would like to purchase their own kit could do so.” |

**Supplemental Table 3: Applicants’ Perspectives on the Utility of Program Websites and Social Media**

| **Online Presence of Residency Programs**  A residency program’s online presence—including social media and websites—greatly impacted applicants’ perception of a program. However, several applicants expressed concerns about the potential consequences of the growing use of social media such as its authenticity in representing programs, the possibility of students being required to utilize social media during the application process, and the increased likelihood that programs would evaluate students by their social media presence. | |
| --- | --- |
| **Sub-Theme** | **Representative Quotations** |
| **Benefits of Strong Websites and Social Media Presence** | - “Programs that put the time into developing an online platform via either social media or a well-developed website were significantly more attractive. It showed that they were willing to innovate and cared to engage applicants during the virtual cycle as best as possible.” - “A strong online presence was key to getting to know a program and being able to reference its information frequently. I likely ranked higher programs with whom I was more engaged online.” - “It was very obvious which programs updated their website and made an effort to create a social media presence in response to the virtual interview season, and this played a huge role in my first impressions of programs!” - “Websites or sessions including details on how the program differs from other programs. What the program chooses to highlight (faculty vs. curriculum vs. resident success, for example) tells a lot about the culture priorities.” |
| **Critiques of Growing Social Media Use** | - “I don't frequent social media and I don't have an Instagram or Twitter. Because of this, I often felt "out of the loop" with programs when I applied. It seemed to be an unwritten rule to check the program's Instagram presence, though without an account myself that was difficult to do. I wished that all of the necessary information was on the public-facing website or intranets that we were provided access to, with only repeat or supplemental info on Instagram or Twitter.” - “I really hope interacting with a program's social media doesn't become a marker of interest or affect ranking. It is by nature very superficial, and plenty of people [who are interested] can't attend Zoom events due to schedule constraints.” - “Social media allows individuals and programs to display what they choose and project an image that may be inconsistent with actuality.” |

**Supplemental Table 4: Applicants’ Perspectives on Assessing Program Compatibility within the Virtual Environment**

| **Evaluation of Program Culture**  While the majority of respondents were satisfied with the virtual Match, several free response comments indicated dissatisfaction and feelings that virtual interviews hindered an applicant’s ability to assess fit with a program. The most commonly reported issue was difficulty in assessing program culture. This included comments remarking on the limited and curated portrayals of programs provided on social media and during virtual interviews, the virtual format not being conducive to meaningful conversations, and the lack of opportunities to observe interactions with hospital personnel. In light of this, it is unsurprising that the most commonly cited component of the application process that was helpful in determining compatibility was casual conversations with residents in small group settings. Many applicants also remarked that their inability to visit the program sites made it more difficult to determine fit and suggested that an in-person second look after programs submit their rank lists could be useful. Students also mentioned that virtual tours and comprehensive information on the area, hospital environment, and diversity were helpful in mitigating this issue. | |
| --- | --- |
| **Sub-Theme** | **Representative Quotations** |
| **Informal Conversations with Residents Were Important for Evaluating Fit** | - “I really appreciated the night-before-the-interview evening sessions with residents. This helped set the tone for the next day and to have something to talk about and ask questions about with the faculty. The best ones were where there were 8+ residents available on Zoom that were limited to 1 hour. The worst were large groups of applicants without any meaningful connection (like a once or twice per application season event) and experimental digital platforms like Gather where residents split up to talk with groups of applicants and we couldn't see them interact with each other.” - “Informal conversations with residents in the waiting rooms helped the most for me—whenever a program faculty member said that I would fit in, I always took it with a grain of salt because I know they are trying to recruit and fill their spots. If a resident said it, however, I took that to heart more because I felt they were really more concerned about finding coworkers that they like to be around.” |
| **Lack of Opportunities to Observe Interactions Between Residents and Other Staff** | - “Seeing how current residents interacted with one another as well as with the PD was a great indicator. Also how the program structured the interview day and the apparent effort they put into making it interesting, engaging, and informational was a big indicator.” - “Evaluating the hospital culture and collegiality among ancillary staff [was a difficulty when evaluating fit]” - “I believe the biggest factor here for me was not being able to see how other hospital/clinical staff acted towards us, the faculty, and current residents. It was impossible to tell if coworkers we'll be working with closely (like nurses, administration, etc.) were going to be welcoming and helpful in our learning experiences versus indifferent or even hostile.” - “The hardest interviews for evaluating "fit" were where only a few (3-4) residents were present and I could not see how they interact as a group; or similarly, when there were many residents but they sent us into breakout rooms where we only spoke to 1-2 residents. It's really important to see the working relationship between the residents.” |
| **Need for Comprehensive Information about Program Environment** | - “As a URM in medicine, I had a really hard time telling if I would fit into the program from the virtual interview since it was such a superficial look at the true culture of the program.” - “Couldn't appreciate the physical structure of the hospital. Maybe do a video of the hospital in general and the department in particular? UPenn's video did just that and it was exceptional. Essentially a resident walked us through the outside of the buildings and then took a tour inside.” |

**Supplemental Table 5: Applicants’ Perspectives on the Future of Residency Interviews**

| **Future of Residency Interviews**  While the majority of respondents supported the continuation of virtual interviews, several comments expressed concerns regarding the possibility of a hybrid interview format. One common concern was that offering virtual interviews as an alternative to in-person interviews would favor applicants who attended in-person and be detrimental to those who may have financial or travel constraints. Many students indicated that rather than providing virtual interviews as an option, programs could create hybrid interview systems by which there is first a virtual interview for screening purposes followed by an in-person interview.  Moreover, despite the majority of respondents agreeing with an interview limit, many students were concerned about implementation of such a policy. Issues voiced included the need for strict enforcement of the cap rather than relying on a honor system and the utility of a universal interview release date. Students also discussed the potential benefits of varying an application cap depending on applicant characteristics. Furthermore, even though most respondents agreed with implementing a preference signaling mechanism, some also voiced concerns regarding students having insufficient information to effectively use tokens and the potential for preference signaling to disturb how the ranking system functions. Notably, many applicants also called for greater transparency on the part of programs regarding how they assess applicant characteristics. | |
| --- | --- |
| **Sub-Theme** | **Representative Quotations** |
| **Equity Concerns with Optional Virtual Interviews** | - “Seems like it needs to be either virtual or in-person, I'd be surprised if programs would take virtual-only applicants as seriously as in-person applicants if they had a mix.” - “I don't think programs should do virtual and in-person interviews. This will stress applicants into thinking about if they should or should not go and will allow people not as interested to just take the virtual interview and not spend money traveling. They should solely do one or the other. It creates disadvantages to the applicants if they do not.” - “I believe virtual interviews and other alternatives to in-person interactions should be further explored as a serious mechanism to augment the traditional methods, rather than as permanent replacements.” - “I think if utilized appropriately, virtual interviews could be an incredibly useful adjunct tool through the residency/fellowship application process. I do believe that it is horrendously difficult, if not impossible, to evaluate "fit" and "culture" of an institution and its individuals virtually. Ideally virtual interviews going forward would be used almost as a pre-screening interview prior to an in-person second look/follow up interview day. I believe virtual interviews are feasible for one-on-one interactions like faculty interviews; however, I think they are absolutely a horrendous platform for group interactions like residency socials as well as incredibly poor at evaluating a physical location (hospital and city).” - “I think the best method is to have virtual interviews for equity purposes, and then optional second looks after programs submit their rank lists but before applicants submit their rank lists.” - “It was really hard and problematic to not see the program, hospital, or city in-person. I feel like virtual interviews encourage hoarding and for programs to screen applicants and only accept who they know.” - “The virtual interview setting has both pros and cons but I believe having it as an option for students could be beneficial going forward, as long as candidates were not weighed differently in various settings.” |
| **Views on Application and Interview Caps** | Mandating Compliance   - “I'm glad the OBGYN national overseeing body suggested a limit for how many programs to apply to. However, I know for a fact that many applicants didn't follow that rule. As a result, the people who followed the recommendation were at a disadvantage. Unfortunately, I think you can't rely on applicants to just "do the right thing," and instead need to mandate the limit.”   Mixed Opinions   - “The caveat to a cap on the number of programs a student may apply to is that it unfairly discriminates against international medical graduates, DO students, and students with low standardized test scores. These students need to apply to a significantly higher number of programs and more than one specialty to get a desirable yield of interviews (sometimes 100-200 programs especially if you're an international grad). A cap on the number of residency interviews is much more feasible and acceptable (again with a caveat of a separate cap for each specialty applied to).” - “Applicants should only be limited in application number if there are criteria met for a certain number of applications. Those with lower scores, are DOs, or are couples matching should be allotted more applications and interviews” - “I think people should only be able to do 20 interviews per specialty to ensure there is no hoarding/wasting programs’ time.” - Though I received very few interview invitations this year, I still do not believe that applicants should be restricted in the number of interviews they may accept. It feels strange to tie the hands of competitive applicants and residency programs to accommodate those who are less competitive. Those applicants made decisions on how to use their time that rendered their applications better than mine, decisions I could have made but did not. They should not be punished for it. I accept the consequences of my own choices.   Universal Interview Release Date   - “All programs within the specialty must release interview invites the same day. This day should theoretically be standardized across all specialties because many students apply to more than one specialty. There could also be a second, third, fourth, etc. rounds of interview invites sent later on in the cycle as interview spots are filled or become available. |
| **Hesitancy Regarding Preference Signaling** | - “Tokens make me nervous, I feel like that would penalize students for going for "reach programs" and would ruin the current rank system. It would return to students ranking based on where they think they will match instead of what they actually want. There are other ways to show you are genuinely interested, reaching out to residents and PDs, showing up prepared for interviews, sending an email that says "you are my top program" to your top, etc.” - “More data would be needed for a token system since applicants might not know how they compare to a program's history of admissions and therefore "waste" some on reaches/safeties.” |
| **Need for Greater Transparency** | - “Program directors blatantly lie to applicants every year about their ranking position with no consequence. This should be strictly forbidden as it is emotionally damaging since applicants are powerless and have way more on the line. The excess of applications is overwhelming programs and my home PD publicly confessed to throwing out apps from anyone she didn't know. This is absurd.” - “The Match is an unjust system that exploits medical students and residents. It is extremely expensive and benefits those with more resources and takes all power away from students due to negating the ability to negotiate contracts. There should be more focus on that rather than virtual or in-person which is only a small part of the overall problem.” - “While I believe limiting applications and interviews is a decent short-term solution, I don't believe this is necessarily an improvement in the long-run. I also believe that something like a "token system" would be detrimental as there were many programs I didn't know enough about until well after applying, and some I didn't know how much I would like until after the completion of the actual interview day. Until significant progress is made on the number and availability of residency positions for all applicants looking into various specialties, the best changes that can be made would be those that make the process more equitable and fair for all.” - “Limiting applications will not help anyone with a "red flag" on their application. The most important thing in limiting applications is program transparency - what are the program's cutoff scores? Are they hard or soft cutoffs? What is their average accepted quartile, or what percentage of their program is historically AOA? Allowing applicants more information to better select which programs are attainable for their individual circumstances will be more efficacious for both applicants and programs because applicants will be better able to assess how they compare to traditionally accepted candidates. Access to more and accurate information would have greatly reduced the number of programs I applied to.” |
