## Supplementary material for "A Match Made in Cyberspace: Applicant Perspectives on a Virtual Residency Application Process": Survey Instrument

### COVID Effects on Residency Applications Survey

The questions in this survey aim to evaluate the student perspective regarding virtual interviews and other effects of the COVID-19 pandemic on the residency application process this cycle. You are being asked to participate in this survey only if you applied to residency programs during the 2020-2021 application season.

The survey should take no longer than 5 minutes to complete. Participation in this survey is voluntary and confidential. By choosing to submit this survey, you are providing consent to be part of this study. You may exit the survey at any time without penalty, and you are free to decline to answer any question. All survey responses will be anonymous and reported only in aggregate.

If you have any questions, please contact Shirley Chen at (Vanderbilt IRB #210572).

#### Demographics

| | |
|---|---|
| Geographic Location of Medical School you Attended | ◯ Alabama<br>◯ Alaska<br>◯ Arizona<br>◯ Arkansas<br>◯ California<br>◯ Colorado<br>◯ Connecticut<br>◯ Delaware<br>◯ Florida<br>◯ Georgia<br>◯ Hawaii<br>◯ Idaho<br>◯ Illinois<br>◯ Indiana<br>◯ Iowa<br>◯ Kansas<br>◯ Kentucky<br>◯ Louisiana<br>◯ Maine<br>◯ Maryland<br>◯ Massachusetts<br>◯ Michigan<br>◯ Minnesota<br>◯ Mississippi<br>◯ Missouri<br>◯ Montana<br>◯ Nebraska<br>◯ Nevada<br>◯ New Hampshire<br>◯ New Jersey<br>◯ New Mexico<br>◯ New York<br>◯ North Carolina<br>◯ North Dakota<br>◯ Ohio<br>◯ Oklahoma<br>◯ Oregon<br>◯ Pennsylvania<br>◯ Rhode Island<br>◯ South Carolina<br>◯ South Dakota<br>◯ Tennessee<br>◯ Texas<br>◯ Utah<br>◯ Vermont<br>◯ Virginia<br>◯ Washington<br>◯ West Virginia<br>◯ Wisconsin<br>◯ Wyoming<br>◯ International (outside of the United States)<br>◯ Prefer not to say |
| Degree Program of Your Medical School | ◯ MD<br>◯ DO<br>◯ MD/PhD<br>◯ DO/PhD<br>◯ MD/MPH<br>◯ DO/MPH<br>◯ MD/MBA<br>◯ DO/MBA<br>◯ Other Dual Degree |

| | |
|---|---|
| Other Dual Degree | __________________________________ |
| Did you match? | ◯ Yes<br>◯ No |
| Where did you rank the program you matched at? | ◯ 1st<br>◯ 2nd<br>◯ 3rd<br>◯ 4th<br>◯ 5th<br>◯ 6th<br>◯ 7th<br>◯ 8th<br>◯ 9th<br>◯ 10th<br>◯ 11th<br>◯ 12th<br>◯ 13th<br>◯ 14th<br>◯ 15th<br>◯ 16th<br>◯ 17th<br>◯ 18th<br>◯ 19th<br>◯ 20th or lower |
| Did you couples match? | ◯ Yes<br>◯ No |
| Were you applying from a transitional year? | ◯ Yes<br>◯ No |
| Please comment on what factors you think most contributed to you not matching. (OPTIONAL) | __________________________________________ |

Specialty(s) that you applied to (Select all that apply). Please exclude any specialties that were only applied to during SOAP.

- ☐ Anesthesiology
- ☐ Child Neurology
- ☐ Dermatology
- ☐ Diagnostic Radiology/Nuclear Medicine
- ☐ Emergency Medicine
- ☐ Emergency Medicine/Anesthesiology
- ☐ Emergency Medicine/Family Medicine
- ☐ Family Medicine
- ☐ Family Medicine/Osteopathic Neuromusculoskeletal Medicine
- ☐ Family Medicine/Preventive Medicine
- ☐ Internal Medicine
- ☐ Internal Medicine/Anesthesiology
- ☐ Internal Medicine/Dermatology
- ☐ Internal Medicine/Emergency Medicine
- ☐ Internal Medicine/Family Practice
- ☐ Internal Medicine/Medical Genetics
- ☐ Internal Medicine/Neurology
- ☐ Internal Medicine/Pediatrics
- ☐ Internal Medicine/Preventive Medicine
- ☐ Internal Medicine/Psychiatry
- ☐ Interventional Radiology
- ☐ Neurodevelopmental Disabilities
- ☐ Neurological Surgery
- ☐ Neurology
- ☐ Nuclear Medicine
- ☐ Obstetrics and Gynecology
- ☐ Ophthalmology
- ☐ Orthopedic Surgery
- ☐ Osteopathic Neuromusculoskeletal Medicine
- ☐ Otolaryngology - Head and Neck Surgery
- ☐ Pathology-Anatomic and Clinical
- ☐ Pediatrics
- ☐ Pediatrics/Anesthesiology
- ☐ Pediatrics/Dermatology
- ☐ Pediatrics/Emergency Medicine
- ☐ Pediatrics/Medical Genetics
- ☐ Pediatrics/Physical Medicine and Rehabilitation
- ☐ Pediatrics/Psychiatry/Child and Adolescent Psychiatry
- ☐ Physical Medicine and Rehabilitation
- ☐ Plastic Surgery
- ☐ Preventive Medicine
- ☐ Psychiatry
- ☐ Psychiatry/Family Practice
- ☐ Psychiatry/Neurology
- ☐ Radiation Oncology
- ☐ Radiology-Diagnostic
- ☐ Surgery
- ☐ Thoracic Surgery
- ☐ Transitional Year
- ☐ Urology
- ☐ Vascular Surgery
- ☐ Other

---

Other Specialty

__________________________________

---

What gender do you most identify with?

- ◯ Male
- ◯ Female
- ◯ Non-binary
- ◯ Other
- ◯ Prefer not to say

| | |
|---|---|
| Other Gender | __________________________________ |
| Race (Select all that apply) | ☐ American Indian/Alaska Native<br>☐ Asian<br>☐ Native Hawaiian/Other Pacific Islander<br>☐ Hispanic/Latino<br>☐ Black/African American<br>☐ White<br>☐ Other<br>☐ Prefer not to say |
| Other Race | __________________________________ |

#### Overall

Please answer the following questions about your experiences with the residency application and interview process this year. The following four questions are for your PREFERRED specialty (i.e. the number one specialty that you hoped to pursue).

| | |
|---|---|
| Number of applications submitted | __________________________________ |
| Number of interview invitations received | __________________________________ |
| Number of interviews attended | __________________________________ |
| Number of programs ranked | __________________________________ |
| Did you apply to more than one specialty? | ◯ Yes<br>◯ No |

Please answer the following questions about your experiences with the residency application and interview process this year. The following four questions are for ANY OTHER specialty you applied to besides your preferred specialty.

| | |
|---|---|
| Number of applications submitted | __________________________________ |
| Number of interview invitations received | __________________________________ |
| Number of interviews attended | __________________________________ |
| Number of programs ranked | __________________________________ |
| Would you have accepted a different number of interviews if they were conducted in person? | ◯ Yes<br>◯ No |

How do you think the number of interviews you accepted would have differed had interviews been conducted in-person?

- ◯ >25% less
- ◯ 11-25% less
- ◯ 1-10% less
- ◯ 1-10% more
- ◯ 11-25% more
- ◯ >25% more

Please explain why you would have accepted a different number of interviews. (OPTIONAL)

__________________________________________

**Please indicate your opinion about the 2020-2021 residency application cycle:**

| | Strongly disagree | Disagree | Neutral | Agree | Strongly agree |
|---|---|---|---|---|---|
| Overall, I am satisfied with my residency application and virtual interview experience this year. | ◯ | ◯ | ◯ | ◯ | ◯ |
| The virtual setting allowed me to effectively evaluate my "fit" within a program. | ◯ | ◯ | ◯ | ◯ | ◯ |
| The virtual setting made the ranking process challenging. | ◯ | ◯ | ◯ | ◯ | ◯ |
| The virtual setting made it difficult to be myself when interacting with programs. | ◯ | ◯ | ◯ | ◯ | ◯ |
| The lack of in-person away rotations limited my ability to assess residency programs | ◯ | ◯ | ◯ | ◯ | ◯ |
| The virtual setting allowed me to adequately assess the cultural competency and/or humility of a program | ◯ | ◯ | ◯ | ◯ | ◯ |
| The virtual setting allowed me to attend more interviews because of reduced financial constraints. | ◯ | ◯ | ◯ | ◯ | ◯ |
| I feel that the virtual setting allowed more students to "hoard" interviews. | ◯ | ◯ | ◯ | ◯ | ◯ |
| I feel that the guidelines published by my preferred specialty's professional society on how many programs to apply to and/or how many interviews to accept were appropriate. | ◯ | ◯ | ◯ | ◯ | ◯ |

| | | | | | |
|---|---|---|---|---|---|
| I feel that the guidance given by my school's advisors about how many programs to apply to and/or how many interviews to accept was appropriate. | ○ | ○ | ○ | ○ | ○ |

#### Technology

**Please characterize your experience with the following social media platforms as used by residency programs during the 2020-2021 residency application cycle.**

| | N/A | Extremely Unhelpful | Unhelpful | Neutral | Helpful | Extremely Helpful |
|---|---|---|---|---|---|---|
| Facebook | ○ | ○ | ○ | ○ | ○ | ○ |
| Instagram | ○ | ○ | ○ | ○ | ○ | ○ |
| LinkedIn | ○ | ○ | ○ | ○ | ○ | ○ |
| Twitter | ○ | ○ | ○ | ○ | ○ | ○ |
| TikTok | ○ | ○ | ○ | ○ | ○ | ○ |

Other Platform(s) ______________________________

Please elaborate on how technology innovations and/or issues impacted your virtual interviews. (OPTIONAL) ______________________________________

#### Evaluating Level of Fit

**Please characterize the overall helpfulness of the following programming methods in allowing you to evaluate your "fit" within a program.**

| | N/A | Extremely Unhelpful | Unhelpful | Neutral | Helpful | Extremely Helpful |
|---|---|---|---|---|---|---|
| Virtual away rotations/information sessions before interview season | ○ | ○ | ○ | ○ | ○ | ○ |
| Virtual social hours before/after interviews | ○ | ○ | ○ | ○ | ○ | ○ |
| Residency open houses | ○ | ○ | ○ | ○ | ○ | ○ |
| Interview day presentations by program directors/department chairs | ○ | ○ | ○ | ○ | ○ | ○ |
| Formal interviews with faculty | ○ | ○ | ○ | ○ | ○ | ○ |
| Formal interviews with residents | ○ | ○ | ○ | ○ | ○ | ○ |
| Informal breakout rooms with residents during interview day | ○ | ○ | ○ | ○ | ○ | ○ |

| | | | | | | |
|---|---|---|---|---|---|---|
| Informal conversations with other applicants outside of interview day | ○ | ○ | ○ | ○ | ○ | ○ |
| Resident/faculty social media accounts | ○ | ○ | ○ | ○ | ○ | ○ |
| Virtual second looks | ○ | ○ | ○ | ○ | ○ | ○ |

Other Programming Method(s) __________________________________

**Please characterize the overall helpfulness of the following components of official residency program websites in allowing you to evaluate your level of "fit" within a program.**

| | N/A | Extremely Unhelpful | Unhelpful | Neutral | Helpful | Extremely Helpful |
|---|---|---|---|---|---|---|
| Virtual tours | ○ | ○ | ○ | ○ | ○ | ○ |
| Resident testimonials | ○ | ○ | ○ | ○ | ○ | ○ |
| Rotation/didactics schedule | ○ | ○ | ○ | ○ | ○ | ○ |
| Call schedule | ○ | ○ | ○ | ○ | ○ | ○ |
| Resident benefits (salary/stipends, insurance, vacation, etc.) | ○ | ○ | ○ | ○ | ○ | ○ |
| Information about the area | ○ | ○ | ○ | ○ | ○ | ○ |
| Graduates' post-residency fellowship/job placement | ○ | ○ | ○ | ○ | ○ | ○ |
| Surgical case data (case numbers/types) | ○ | ○ | ○ | ○ | ○ | ○ |
| Current residents' bios/photos | ○ | ○ | ○ | ○ | ○ | ○ |
| Faculty bios/photos | ○ | ○ | ○ | ○ | ○ | ○ |

Other Website Component(s) __________________________________

Please elaborate on components of this year's residency application and interview process that were most helpful in allowing you to evaluate "fit" with a program. (OPTIONAL) __________________________________________

Please describe the difficulties you faced with evaluating your level of "fit" within a program in the virtual setting. (OPTIONAL) __________________________________________

#### Resources

**Please indicate your overall thoughts about availability of resources during the 2020-2021 residency application cycle.**

| | N/A | Strongly Disagree | Disagree | Neutral | Agree | Strongly Agree |
|---|---|---|---|---|---|---|
| Finances were a concern for me when applying to residency. | ○ | ○ | ○ | ○ | ○ | ○ |
| I took out (a) loan(s) to complete my residency applications. | ○ | ○ | ○ | ○ | ○ | ○ |
| I felt that I had the resources to successfully complete my residency interviews. | ○ | ○ | ○ | ○ | ○ | ○ |
| My school provided adequate physical space for virtual interviews. | ○ | ○ | ○ | ○ | ○ | ○ |
| My school provided adequate financial assistance for virtual interviews. | ○ | ○ | ○ | ○ | ○ | ○ |
| My school provided adequate electronic equipment (webcams, lights, etc.) for virtual interviews. | ○ | ○ | ○ | ○ | ○ | ○ |
| My school provided adequate coaching and/or guidance on how to prepare for virtual interviews. | ○ | ○ | ○ | ○ | ○ | ○ |

Please elaborate on resources that your school provided which were helpful when preparing for residency applications and virtual interviews. (OPTIONAL) ________________________________

#### Future Cycles

**Please indicate your opinion about future residency application cycles.**

| | N/A | Strongly Disagree | Disagree | Neutral | Agree | Strongly Agree |
|---|---|---|---|---|---|---|
| The virtual setting increases financial access to interviews. | ○ | ○ | ○ | ○ | ○ | ○ |
| The virtual setting decreases an applicant's ability to show genuine interest in a program. | ○ | ○ | ○ | ○ | ○ | ○ |

| | | | | | | |
|---|---|---|---|---|---|---|
| Applicants should only be permitted to APPLY to a limited number of programs. | ○ | ○ | ○ | ○ | ○ | ○ |
| Applicants should only be permitted to INTERVIEW at a limited number of programs. | ○ | ○ | ○ | ○ | ○ | ○ |
| Applicants should have a mechanism to indicate genuine interest in a program, commonly referred to as a "token system". | ○ | ○ | ○ | ○ | ○ | ○ |
| Residency programs should continue to provide virtual interviews as an option even when in-person interviews are offered. | ○ | ○ | ○ | ○ | ○ | ○ |

#### Concluding Thoughts

Please share anything else you think is important about your residency application and virtual interview experience this year. (OPTIONAL) __________________________________
