## Supplementary material for "A Match Made in Cyberspace: Applicant Perspectives on a Virtual Residency Application Process": Figure 1

**Figure 1: Recommendations for medical schools and residency programs in a virtual Match setting**

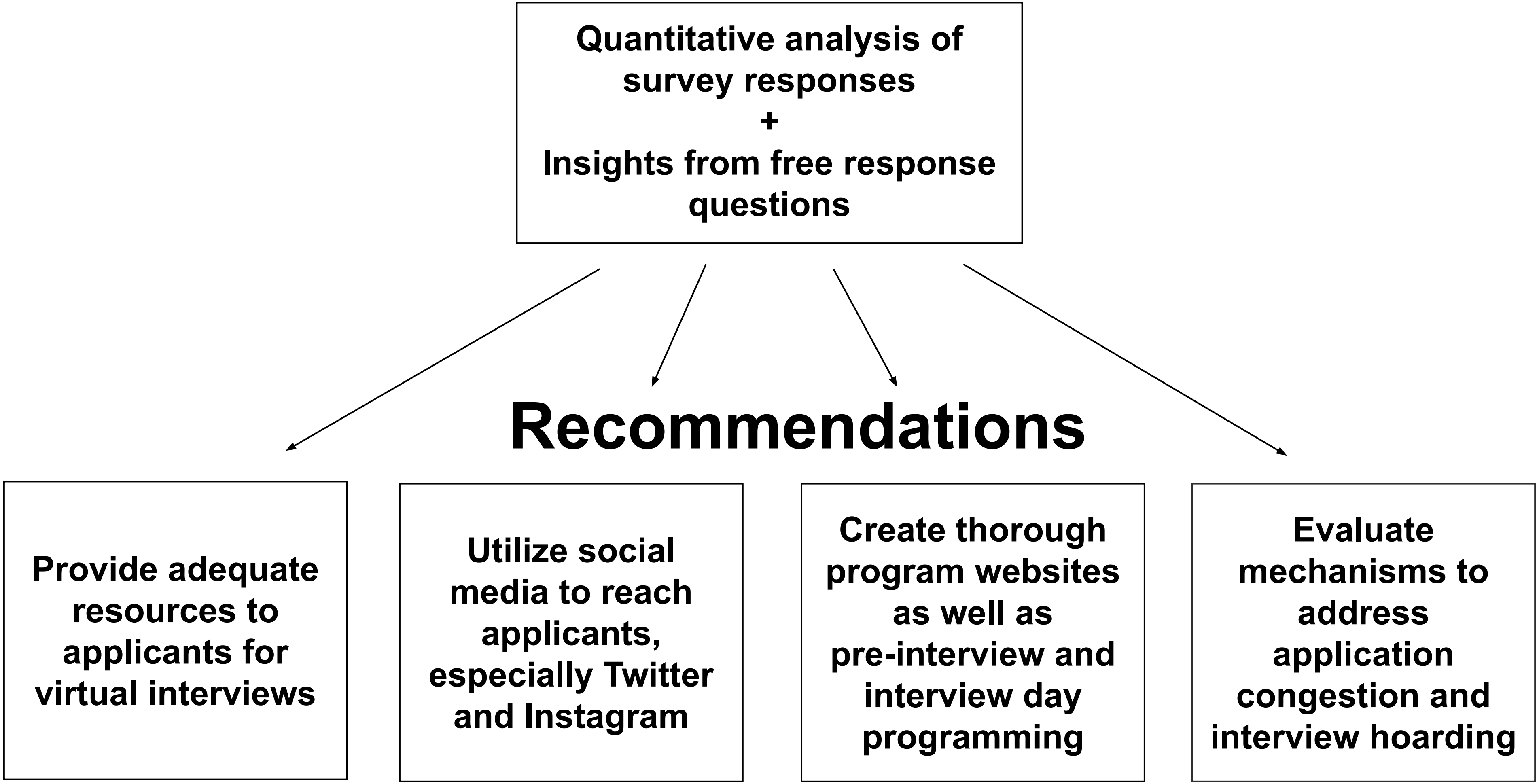
